# Creating safe spaces to improve urban adolescent health and family planning outcomes: lessons from a multi-sectoral intervention in Nigeria

**DOI:** 10.1101/2025.08.26.25334416

**Authors:** Babajide O. Daini, Mojisola O. Alere, Chukwudike Akanegbu, Babatunde Odusolu, Boladale Akin-Kolapo, Gertrude Odezugo

## Abstract

Out-of-school adolescents (OSY) in urban slums face layered vulnerabilities and are often excluded from mainstream reproductive health services. The Youth-Powered Ecosystem to Advance Urban Adolescent Health (YPE4AH) project in Nigeria implemented a multi-sectoral, safe-space-centered model to improve access to family planning (FP) for this group. The project, implemented across Lagos and Kano States from 2021 to 2024, utilized a hub-and-spoke model, combining youth-designed safe spaces (Youth Hubs) with a network of adolescent-friendly health providers. All participants received a foundational life-skills curriculum (SKILLZ), a financial literacy training (SKILLZ Club), and supplemental livelihood and leadership tracks, driven by peer-learning and mentoring. The design utilized a mixed-method approach comprising review of service statistics, longitudinal survey from program participants, and regression analysis to assess trends in uptake of FP services. Quantitative data were drawn from service statistics, longitudinal survey data, and regression analysis of FP uptake. Over 66,000 adolescents completed the SKILLZ curriculum, with a 90% transition into SKILLZ Club. Adolescent preference for non-traditional health spaces was validated, with hubs acting as entry points to broader health services. More than 75,000 adolescents accessed FP services through hub-based and referral pathways, contributing 16,303 couple years of protection (CYPs) and averting an estimated 7,324 unintended pregnancies and 39 maternal deaths. FP uptake increased steadily over time, with a diverse method mix and regional variation in preferences. Integration was achieved through collaboration with public systems, not by co-locating services in primary healthcare centers, but by embedding free FP commodities and referral systems within adolescent-friendly environments. For OSY (especially those in urban slums), safe spaces grounded in youth participation and community trust can serve as critical platforms for FP access. When designed as entry points (not endpoints), and supported by intentional integration with government supply systems, such models offer an alternative pathway to adolescent-responsive health delivery. The YPE4AH experience challenges traditional models of integration and provides a replicable blueprint for reaching marginalized adolescents at scale.

## Introduction

Adolescent sexual and reproductive health (SRH) remains a critical area of concern globally, particularly in low and middle-income countries where access to comprehensive SRH services is limited. In sub-Saharan Africa, this challenge is compounded by structural inequalities, cultural norms, and systemic gaps in youth-friendly service delivery [1,2].

Nigeria, the most populous country in Africa, presents a striking example of these dynamics. Its demographic structure reflects a growing youthful population, with a broad-based population pyramid that underscores the urgency of addressing adolescent health needs [3,4,5]. Despite efforts to improve access, many adolescents remain unable to access contraceptives when needed [5]. This gap often results in unintended pregnancies, which, in most instances, end in unsafe abortions. Consequently, the utilization of reproductive health (RH) services has been identified as a critical step toward improving maternal health [6,7], an imperative aligned with Sustainable Development Goals (SDGs) 3 and 5.

Compounding the challenges posed by a rapidly growing youth population is Nigeria’s persistent out-of-school crisis [8]. The country hosts the largest number of out-of-school children globally, with estimates suggesting that one in every five out-of-school children in the world resides in Nigeria [8,9]. Out-of-school adolescents (OSY), particularly those living in urban informal settlements, constitute one of the most underserved populations in SRH programming [10,11]. Their exclusion from formal structures (schools and conventional health systems) places them at a significant disadvantage, exposing them to heightened risks and limited support [11].

To address the reproductive health needs of adolescents, Nigeria has made commendable strides through policy and programmatic initiatives. These include the development of a national policy on the health and development of young people in Nigeria [12], the formulation of an action plan to guide the integration of adolescent health services into the primary health care (PHC) system [13,14], and the establishment of National Standards and a Minimum Service Package for Adolescent and Youth-Friendly Health Services [15]. Several adolescent-focused interventions have been implemented based on this policy direction, many of which are tailored to in-school adolescents through platforms such as the Family Life and HIV/AIDS Education (FLHE) curriculum at the basic and secondary education levels [16].

However, while attention to school-based interventions and PHC integration has grown, OSY, especially those residing in urban slums, often fall through systemic cracks. They face layered vulnerabilities, including disrupted education, economic precarity, early sexual debut, and limited access to accurate SRH information [9]. In a context where adolescent fertility remains high and unmet need for contraception persists, the exclusion of OSY from structured services continues to fuel preventable risks such as unintended pregnancies, unsafe abortions, and maternal mortality [10,17].

Traditional service delivery models (whether facility-based or school-linked) frequently miss OSY due to barriers such as physical inaccessibility, rigid operating hours, perceived judgment from health workers, and lack of confidentiality. These challenges are further exacerbated by social stigma, misinformation, and limited decision-making autonomy, and thus necessitate revised approaches for OSY [18,19].

Recognizing these service delivery gaps, USAID co-designed the Youth-Powered Ecosystem to Advance Urban Adolescent Health (YPE4AH) project to create a supportive platform for OSY in urban slums, positioning family planning (FP) as a gateway to broader adolescent health and empowerment. Implemented across Lagos and Kano States from 2021 to 2024, YPE4AH introduced youth-designed safe spaces (“Youth Hubs”), foundational life skills education, and a network of trained, adolescent-friendly providers. The model builds on existing guidance proposed for adolescent safe spaces [20], and aimed to shift both the where and how of adolescent health engagement, building environments that were proximate, relational, and youth-owned.

This paper documents the FP-specific outcomes of YPE4AH, examining whether safe spaces, supported by peer mentorship and integrated curricula, can serve as effective entry points for contraceptive uptake and continued use among OSY living in urban slums.

### Program Description

YPE4AH was implemented in urban informal communities in Lagos and Kano States, targeting out-of-school adolescents (OSY) aged 15 to 19 years. The project was designed to address multiple barriers to FP access by embedding services within a youth-led ecosystem of support, anchored in safe, adolescent-centered environments. Central to the program were TEENSMATA Youth Hubs; physically safe spaces where adolescents could gather, learn, access FP information and services, and be referred to other facilities as needed. Collaborations with state governments in Lagos and Kano ensured that the Youth Hubs were hosted as stand-alone spaces within established government structures, but not integrated within traditional PHCs where some other adolescent programming have utilized.

The Youth Hubs served as the focal points of a hub-and-spoke delivery model. Each hub was linked to a network of adolescent-friendly providers at surrounding spoke facilities, including primary health centers (PHCs), private hospitals, community pharmacies, and patent medicine vendors (PPMVs). Through the government collaboration established, free FP commodities were also provided for the hubs where they could be accessed by adolescents and reported back through the state’s supply chain working through nearby PHCs. This supply of commodities was the major source of FP commodities for the YPE4AH project through the implementation period.

Health providers were trained in youth-friendly service delivery, and referral linkages ensured that services unavailable at the Youth Hubs could be accessed nearby. The implementation incorporated a delivery of FP/RH services to adolescents that incorporates key standards from the nine standards of the Nigeria National Standards & Minimum Service Package for Adolescent &Youth-Friendly Health Services[15], shown in Table 1 below.

**Table 1:**
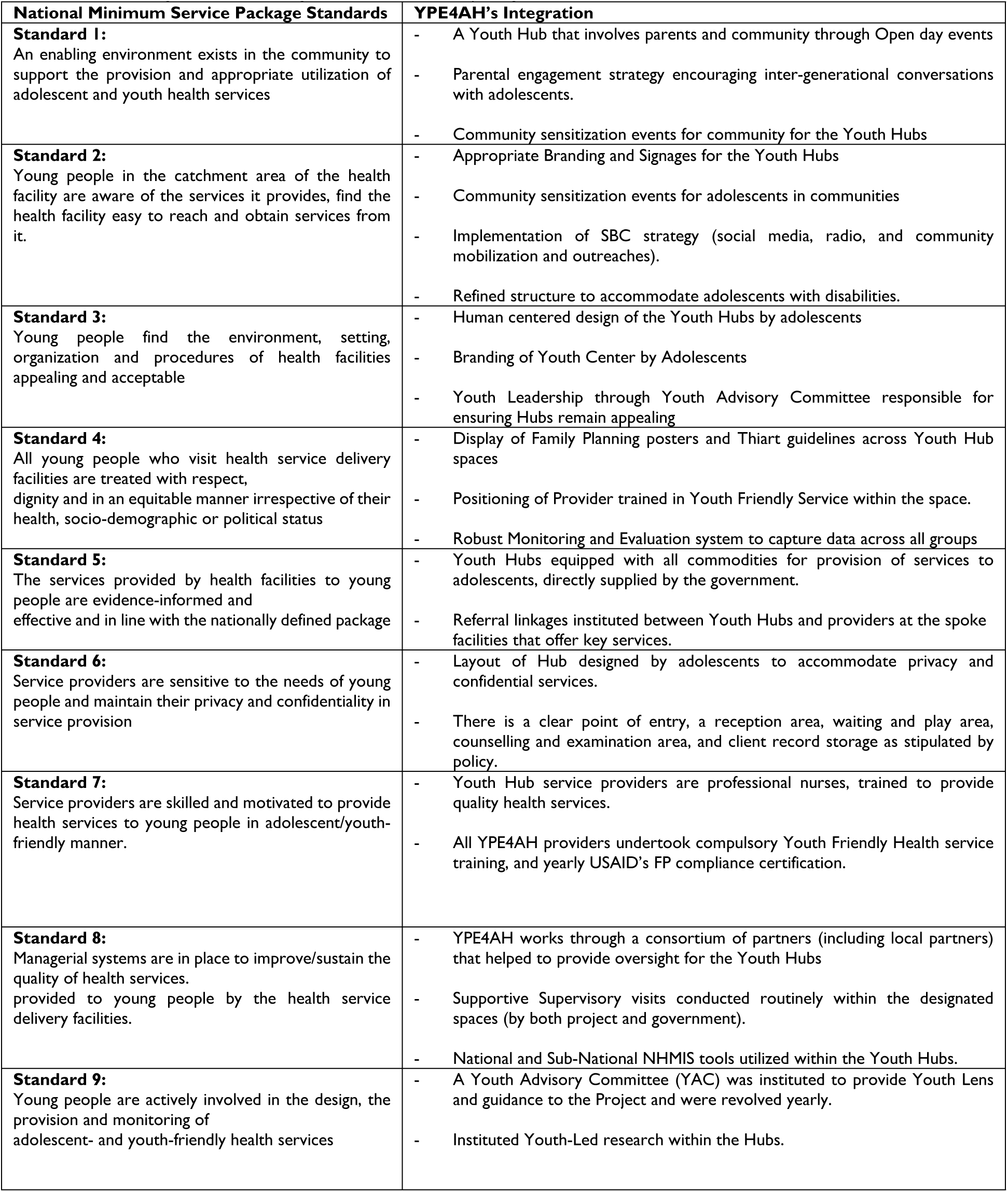
YPE4AH’s Integration with the Nigeria’s Minimum service package Standards for Adolescents.

All participating adolescents received a foundational life-skills curriculum, SKILLZ, adapted from Grassroot Soccer [21,22]. This play-based, group learning intervention used sports metaphors and activities to build knowledge and skills related to health, gender, relationships, and goal-setting, and ran over an initial 2-week period for each cohort. Lagos and Kano states presented two broad scenarios in terms of reaching adolescents with the SKILLZ intervention. In Lagos, the approach was built around a mixed-sex curriculum, comprising of both female and male adolescents in a 60-40 ratio, whereas in Kano, a single-sex curriculum was deployed for three distinct groups of adolescents unmarried boys, unmarried girls, and married girls) in a 40-30-30 ratio. This was based on the formative research that showed that a large proportion of adolescents were already married, unlike in those in Lagos. These curricula were distinct for each group, with major differences determined by socio-cultural and religious context for the groups. Adolescents who completed SKILLZ were eligible to proceed to SKILLZ Club, which offered additional modules focused on financial literacy, entrepreneurship, and career readiness.

The project also integrated other youth development elements, such as mentorship by near-peer coaches, adolescent-led governance through Youth Advisory Committees, and community engagement to build support for adolescent SRH access. Although the primary focus was on FP, the YPE4AH model emphasized the importance of addressing the broader determinants of adolescent well-being [Figure 1].

**Figure 1:**
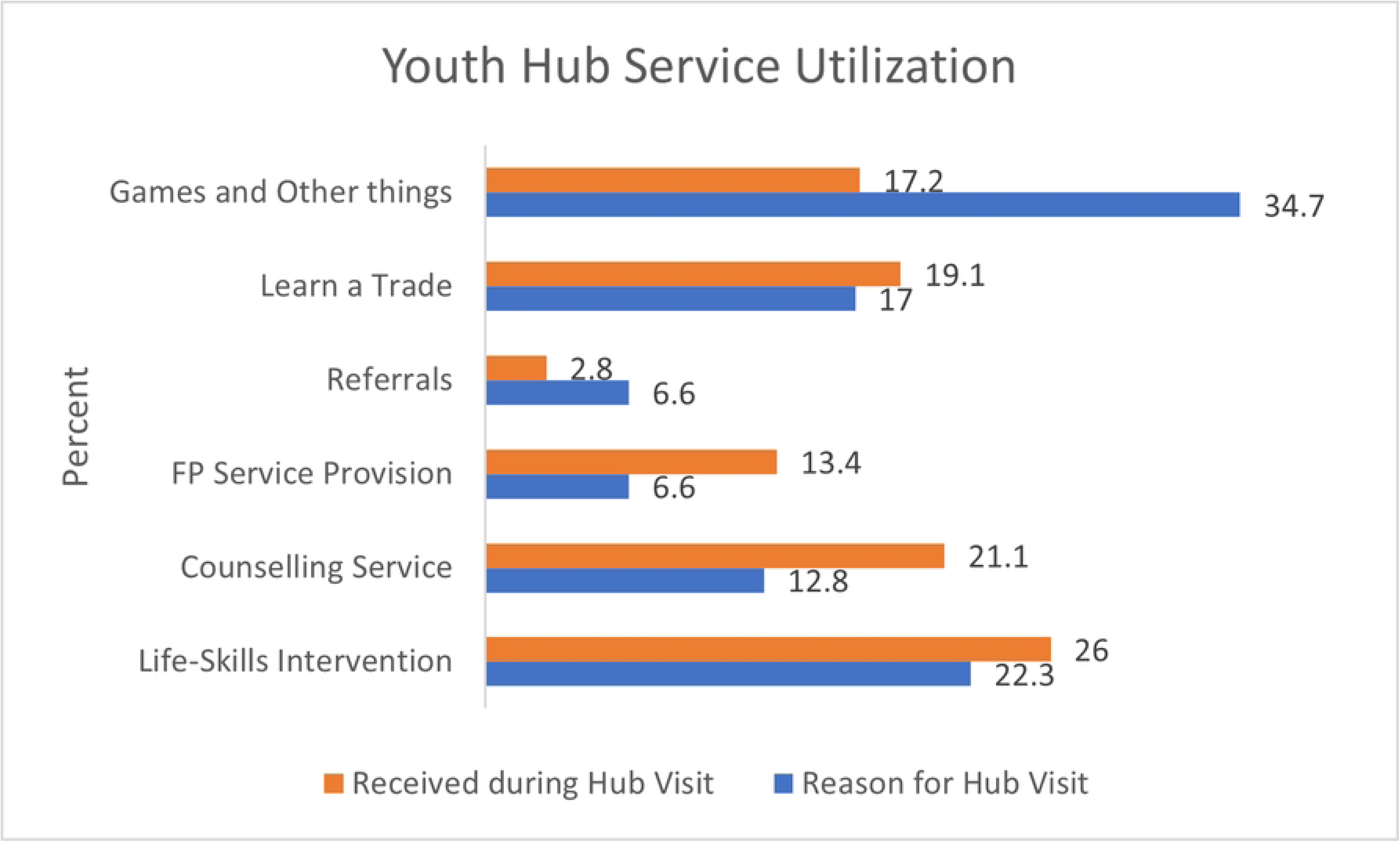
Linking YPE4AH Objectives with existing Adolescent Frameworks

The holistic approach to adolescent well-being is in line with current best practice about adolescent health, as it addresses all domains in theorized adolescent well-being frameworks that work [23,24].

## Methods

This study draws from implementation data collected between June 2021 and April 2024 across Lagos and Kano States. The design utilized a mixed-method approach comprising review of service statistics, longitudinal survey from program participants, and regression analysis to assess trends in uptake of FP services.

Service statistics were collected across all Youth Hubs and linked spoke facilities using standardized family planning (FP) registers and digital data tools (on project’s database), accessed in May 2024. These data captured FP counseling, method initiation, revisits, and referral patterns. All data from the secondary analysis of registers were anonymized and did not contain any personally identifiable information (PII). Concurrently, a longitudinal survey was implemented with adolescents enrolled in the program who completed the SKILLZ training, defined as attending ≥80% of sessions. FP data were reported as new acceptors (individuals who had never used FP prior to the visit or who had lapsed at their last sexual intercourse) and revisits (continued users, irrespective of provider). Baseline information was obtained from enrollment forms and pre-assessments at program commencement, and follow-up assessments were conducted one-year post-intervention by trained data collectors (starting September 2022). At follow-up, an additional consent process was undertaken to confirm continued participation. Ethical approval for the longitudinal survey was obtained from the National Health Research Ethics Committee of Nigeria (NHREC/01/01/2007-05/09/2022).

Trend analysis and regression models were applied to monthly service data to assess changes in counseling and method uptake over time. The Impact 2 model [25] was used to estimate couple years of protection (CYP), unintended pregnancies averted, unsafe abortions, and maternal deaths. The focus of this paper is on FP-specific outcomes and behavior change; broader outcomes related to leadership, livelihoods, or long-term socio-economic status were not assessed due to inconsistent exposure across participants.

To reduce bias, standardized FP registers and digital data tools were used across all hubs and spoke facilities to ensure consistency of reporting. Uniform operational definitions were applied (e.g., for new acceptors and revisits) to limit misclassification and calculations of FP outcomes.

For the longitudinal survey, retention bias was addressed by following the same cohort of adolescents over time with active re-consent at follow-up. In addition, all secondary service data were anonymized prior to analysis, preventing identification and minimizing reporting bias linked to confidentiality concerns.

Efforts to minimize loss to follow-up in the longitudinal survey included maintaining contact with participants through the Youth Hubs, which remained functional within the implementing communities. Nevertheless, some attrition occurred over the one-year interval, as adolescents returned to school, relocated, or changed the contact details initially provided. The analysis was restricted to respondents with both baseline and follow-up data, with frequencies reported only for this cohort.

## Results

### Utilization of Safe Spaces by Out-of-School Youth (OSY)

From project inception in 2021 to April 2024, the YPE4AH project reached a total of 66,668 adolescents with the SKILLZ life-skills curriculum, as shown in Table 2. Participation was relatively balanced between Lagos and Kano states, with 56.7% identifying as female, and 59.3% aged 15–17 years. Majority of OSY engagements held within the communities (97.5%), especially where adolescents could be found within the community. Approximately 11% were married at the time of enrollment, highlighting the heightened SRH vulnerability among OSY participants.

**Table 2:** Characteristics of OSY Reached by YPE4AH Project.

| Characteristics | Lagos<br>% (N) | Kano<br>% (N) | Total<br>% (N) |
| --- | --- | --- | --- |
| **STATE** |  |  |  |
| Female | 21,624 | 16,157 | 56.7 (37,781) |
| Male | 17,940 | 10,947 | 43.3 (28,887) |
| **MARITAL STATUS** |  |  |  |
| Single | 39,488 | 19,611 | 88.6 (59,099) |
| Married | 49 | 7,491 | 11.3 (7,540) |
| Previously Married | 27 | 2 | 0.04 (29) |
| **AGE** |  |  |  |
| 15-17 Years | 26,050 | 13,488 | 59.3 (39,538) |
| 18-19 Years | 13,514 | 13,616 | 40.7 (27,130) |
| **DELIVERY METHOD** |  |  |  |
| Youth Hub | 18,735 | 11,145 | 44.8 (29,883) |
| Community Based | 19,714 | 15,431 | 52.7 (35,145) |
| Youth Camp | 1,115 | 525 | 2.5 (1,640) |
| **ENROLLED IN SKILLZ CLUB** |  |  |  |
| Yes | 33,499 | 23,639 | 85.7 (57,138) |
| No | 6,065 | 3,465 | 14.3 (9,530) |
| **TOTAL** | **59.3 (39,564)** | **40.7 (27,104)** | **100 (66,668)** |

Notably, nearly 90% of SKILLZ graduates transitioned to the optional SKILLZ Club focused on financial literacy. Longitudinal tracking revealed that 55% of adolescents continued to visit the Youth Hubs weekly one year after program exposure, reinforcing the safe spaces’ relevance in adolescents’ daily routines.

Initial formative research emphasized OSYs’ desire for confidential, safe environments with relatable adults and peer mentors. These expectations were met through the creation of six Youth Hubs across Lagos and Kano. Adolescents cited the hubs’ informal nature, featuring games, music, and youth-led activities, as central to their continued use. While many initially came to socialize, the relaxed atmosphere created soft-entry pathways to health services. Figure 2 shows utilization of the safe spaces by OSY and how these adolescents sought services while at the Youth Hubs.

**Figure 2:**
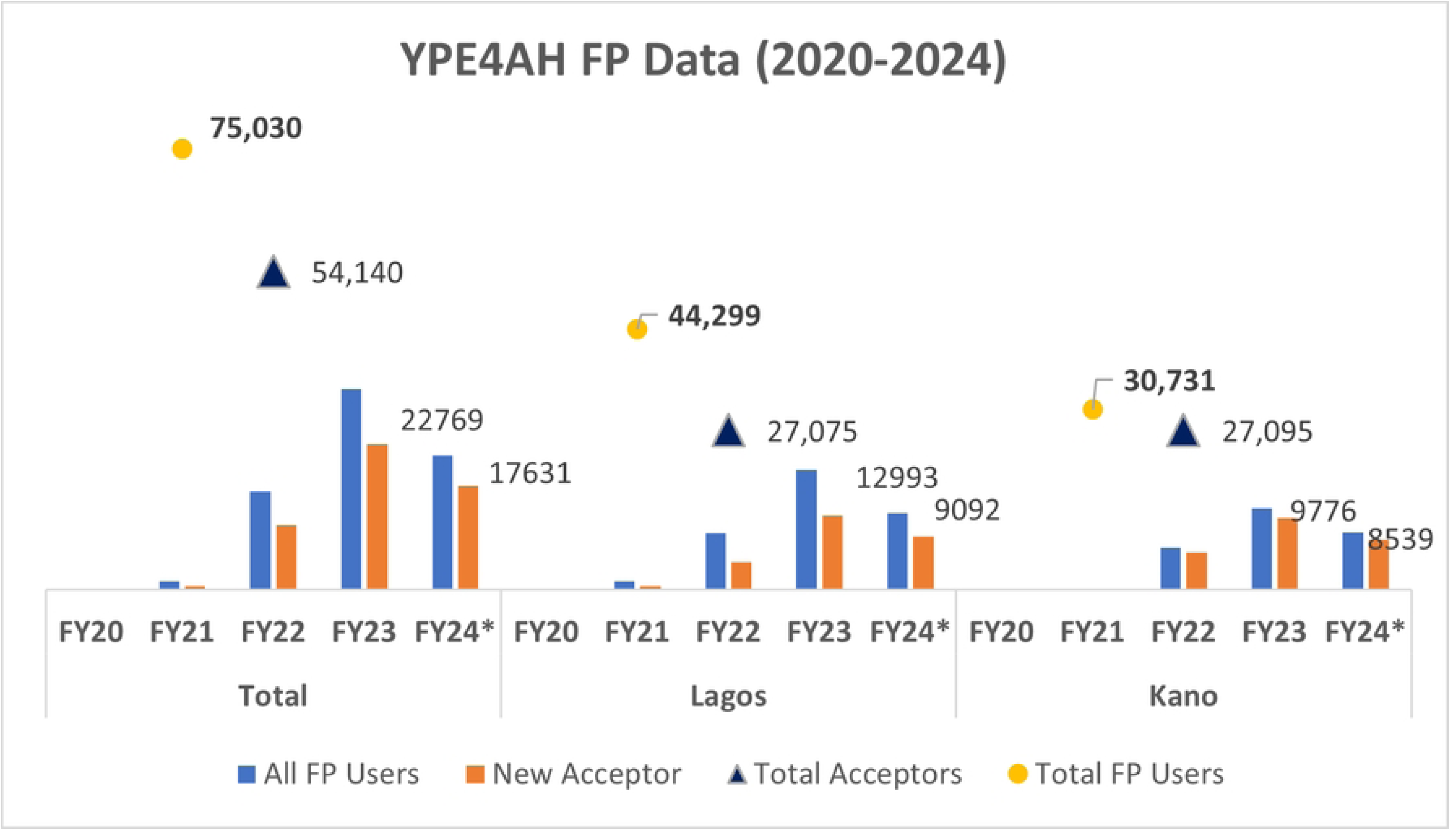
Adolescents’ Utilization of Youth Hubs

By design, the Hubs operated outside traditional PHCs to address stigma, lack of privacy, and distrust often associated with institutional care. This proved critical. Feedback consistently pointed to adolescents’ preference for “non-adult” spaces. The Youth Hubs thus emerged as adolescent-accepted alternatives, particularly for those reluctant to seek care elsewhere.

### Youth-Friendly Health Services at the Safe Spaces

Between June 2021 and March 2024, the Youth Hubs recorded 39,540 FP counseling sessions with adolescents, of which 28,347 led to FP method uptake. Counseling sessions often preceded method adoption, particularly among female adolescents who expressed a need for deliberation. In contrast, male users frequently requested condoms during their first visits.

The SKILLZ curriculum, which included comprehensive SRH content, contributed to increased knowledge and positive attitude shifts. Findings comparing pre- and post-evaluations of the SKILLZ training revealed that 70 percent of these adolescents increased gender equitable beliefs directly as a result of the interventions. There was also a remarkable increase in perception about key FP related concepts; with more changes among those from Kano (a conservative state in Northern Nigeria). The sessions resulted in increased knowledge about FP and specific methods, where to access the services, and intention to use FP in the future (which could be immediately during the interventions or whenever adolescents were ready) (see Figure 3).

**Figure 3:**
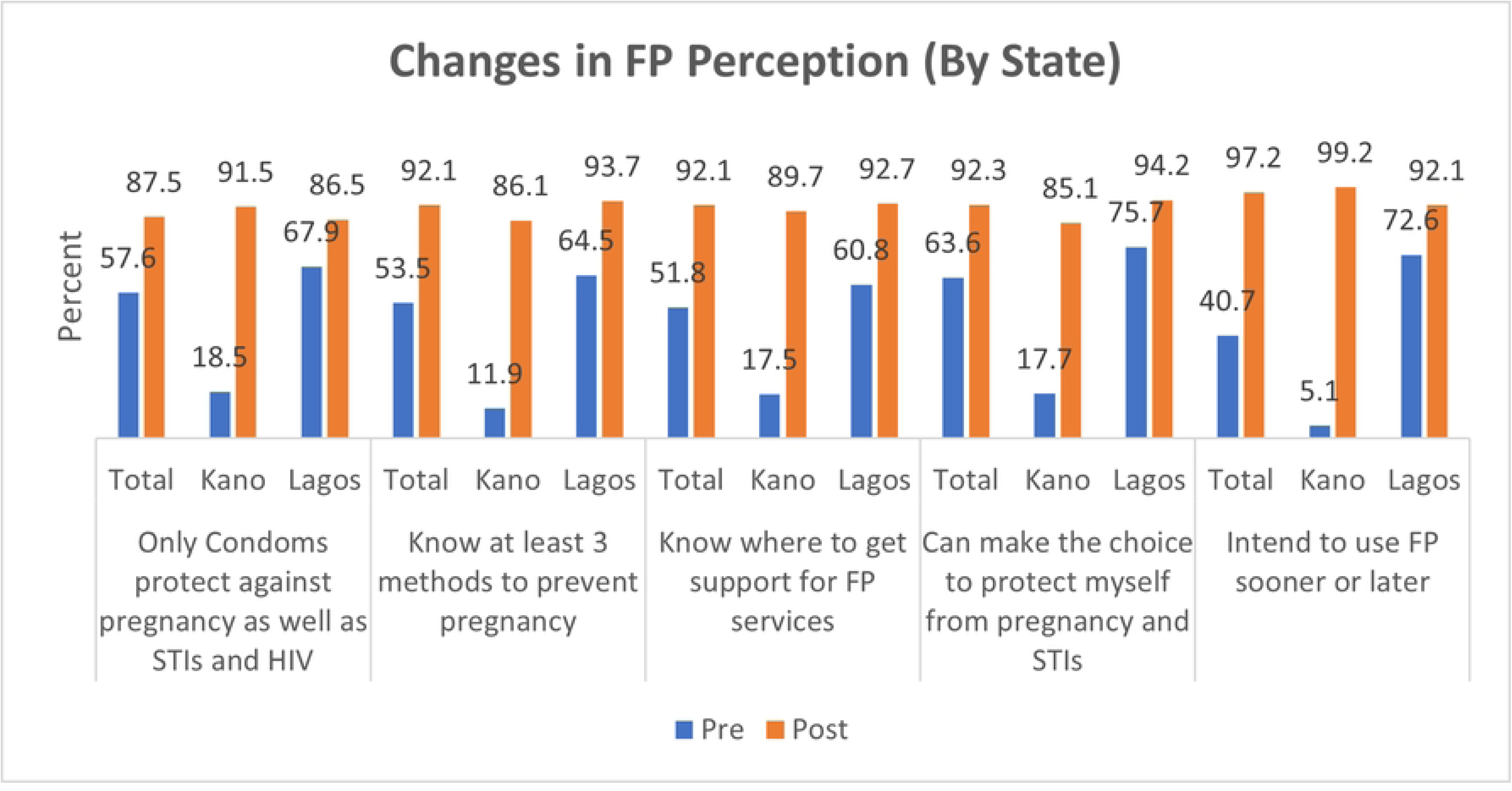
Changes in FP/RH Perception following SKILLZ Intervention (by State)

FP discussions within the curriculum and during peer-led sessions normalized contraceptive use. Adolescents described the hubs as places where “you can ask questions without shame,” and credited peer coaches with dispelling myths and improving comfort with method use.

The project’s hub-and-spoke model allowed for bi-directional referrals between Youth Hubs and 246 trained health providers, ensuring access to services not available directly on-site, such as IUCD insertion and advanced care for GBV survivors. About 150 adolescents were referred from Youth Hubs for IUCD services.

### Assessing Family Planning Outcomes

Within the Youth Hubs, by April 2024, a total of 15,858 adolescents who completed the SKILLZ curriculum adopted a modern FP method. This translates to a 24% conversion rate, a significant jump from the 7.7% reported among a comparable cohort in the 2020 formative research. The contraceptive method-mix included injectables and implants as most preferred, especially in Kano where 50% of all methods were injectables.

Regression analysis of quarterly data from the safe space from June 2021 to March 2024 showed statistically significant trends, with FP counseling increased by 558 adolescents per quarter (R² = 0.82, *p* < 0.001), FP uptake increased by 468 per quarter (R² = 0.85, *p* **<** 0.001), and Conversion from counseling to uptake increased by 3% per month (R² = 0.36, *p* = 0.005).

Beyond direct service uptake in the Hubs, the program’s broader service ecosystem (including referral linkages to PHCs, private clinics, and pharmacies) delivered FP services to 75,030 adolescents between June 2021 and April 2024. This included clients who were referred to spoke facilities from the hubs, as well as those reached through provider outreach, youth events, and community mobilization. Among the 75,030 total FP users, more than 70% were first-time users, underscoring the program’s effectiveness in reaching new adopters. Figure 4 illustrates the upward trend in total FP users across quarters, disaggregated by source.

**Figure 4:**
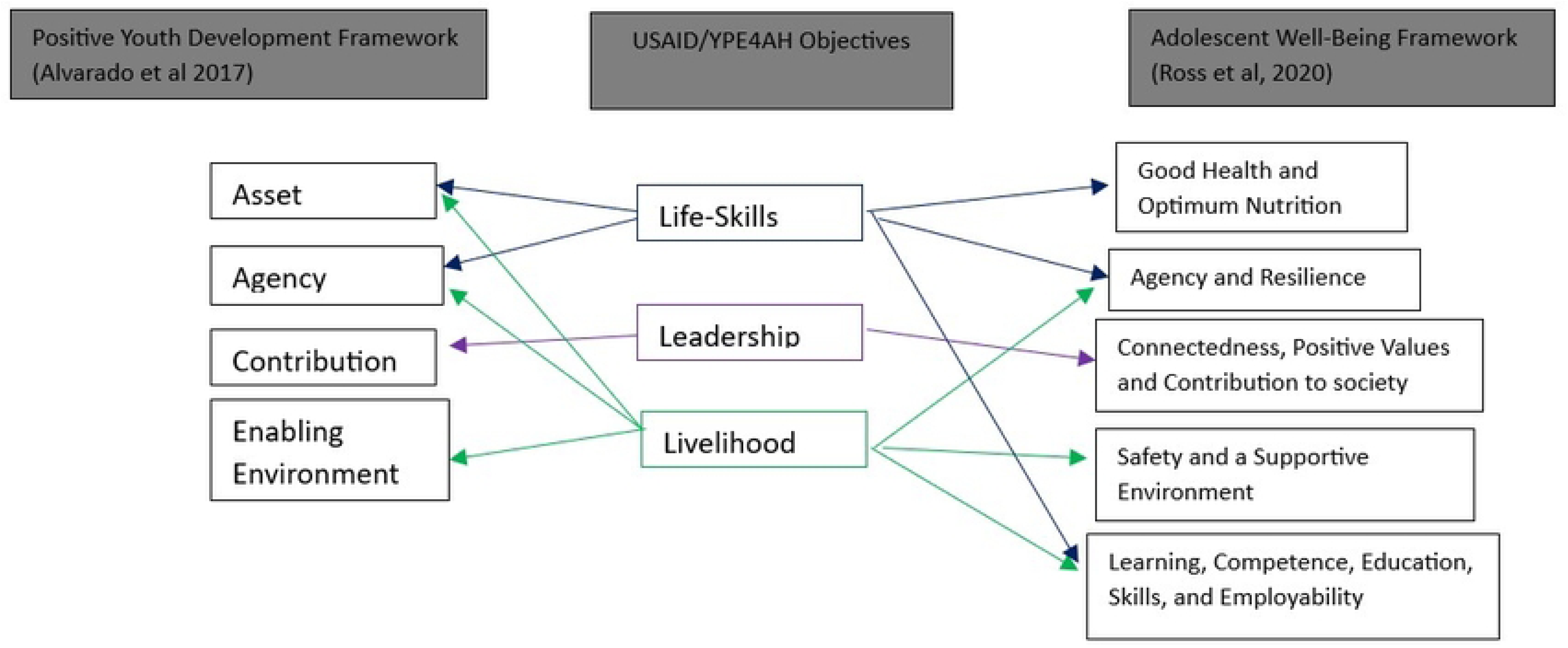
FP Service Data from YPE4AHs (2020-2024)

Impact modeling using MSI’s Impact 2 tool estimated that the program’s FP services generated 16,303 couple years of protection (CYPs), averted 7,324 unintended pregnancies, 2,995 unsafe abortions, and 39 maternal deaths.

Longitudinal follow-up revealed sustained behavior change: one-year post-intervention, current use of FP among sexually active adolescents increased from 18.5% to 27.8%. Table 3 compares these changes with baseline figures. Ever use of contraception rose from 7.7% to 30.8%, and among sexually active youth, contraceptive use at the time of the survey reached 69.6%.

**Table 3:**
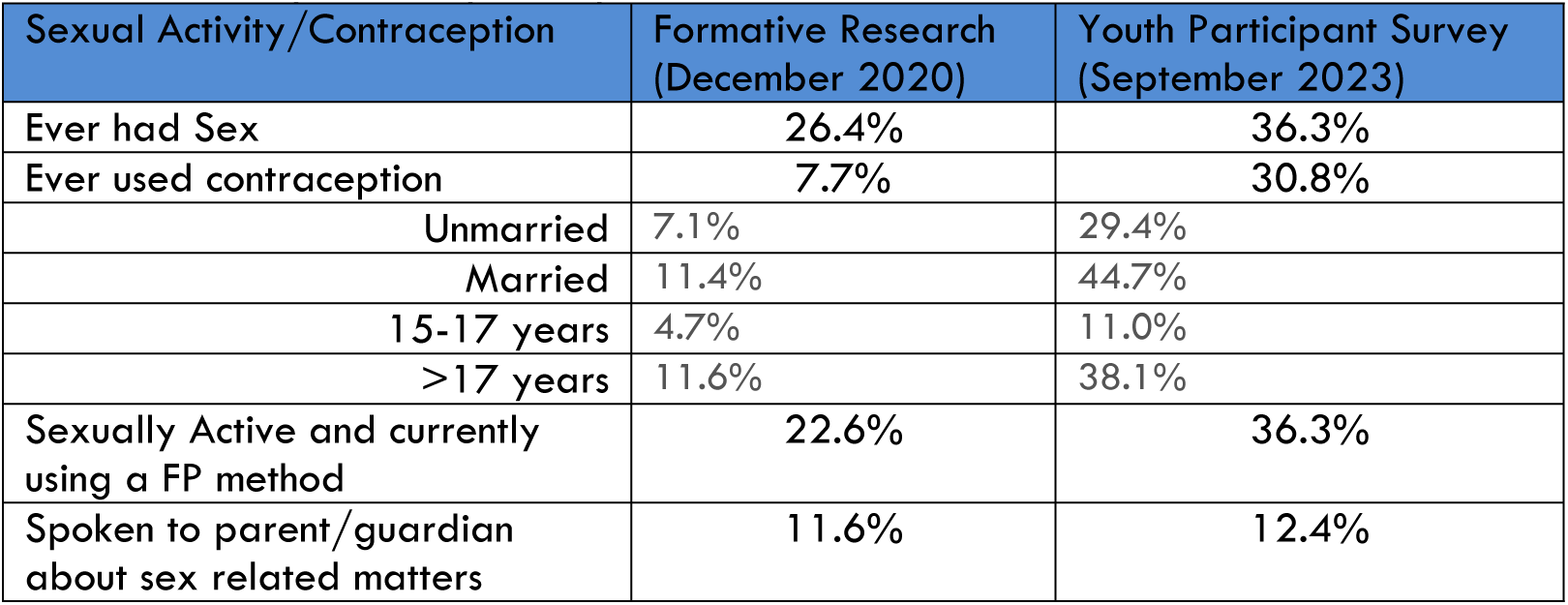
Comparing FP Findings among adolescent cohorts with Formative Research.

Improvements in intergenerational dialogue were also noted, albeit modest. The proportion of adolescents reporting having spoken to a parent or guardian about sex increased from 11.6% to 12.4% between the formative research and follow-up in September 2023.

## Discussion

The findings from the YPE4AH project underscore the potential of safe-space-centered models to improve FP access among out-of-school adolescents (OSY) in urban slums. The program not only achieved significant reach and uptake, but also shifted how and where adolescents seek reproductive health services. The evidence affirms that adolescent-led, community-based platforms, when well-structured and trusted, can serve as critical anchors for SRH interventions.

Over 66,000 adolescents participated in the foundational SKILLZ curriculum, and almost nine out of them participants transitioned to SKILLZ Club, underscoring the relevance and attractiveness of the safe space approach. The design of Youth Hubs as informal, adolescent-owned platforms offering games, music, life skills education, and peer mentoring allowed OSY (many of whom experience high levels of marginalization) to find entry points that felt safe and appropriate, within their immediate communities. In many cases OSYs interacted with the project before ever visiting the Youth Hub space, as the hubs through near-peers moved rotationally within their communities. Nearly six of every 10 participants visited hubs more than once, reinforcing the value of repeated exposure in building trust and shifting attitudes toward SRH services.

This approach challenges earlier findings from Villarruel and colleagues [26], who noted that life-skills interventions often led to short-term behavioral changes without consistent impact on contraceptive use. In contrast, YPE4AH’s embedded hub model provided OSY the opportunity to engage over time within their own communities, enabling gradual but sustained shifts. Similarly, Adesola [4] and Brieger [18] had previously emphasized the difficulty of using peer education alone to reach OSY, recommending complementary interventions to address the structural vulnerabilities this group faces. The YPE4AH program met this recommendation by embedding SRH services within a broader youth development agenda (including financial literacy, livelihood, and leadership) thus creating a more enabling environment.

Furthermore, Erulkal’s framing, that “if we design well to meet the needs of OSY, we can have positive impact even in difficult places” [27], is exemplified in YPE4AH’s northern Nigeria implementation. Despite restrictive social norms, over 11% of enrollees were married adolescents, and many other unmarried adolescents accessed FP services for the first time. This success suggests that adolescent-designed, community-based spaces can overcome even deeply entrenched barriers when trust and relevance are prioritized.

The YPE4AH model also demonstrated that adolescent-friendly care does not have to be confined to PHCs or hospital environments. By training a diverse network of providers (including community pharmacies and patent medicine vendors), and collaborating with the government to secure free FP commodities within the safe space, the project affirmed that proximity, privacy, and relational trust matter as much as clinical infrastructure. This is particularly important for OSY, who may not feel comfortable in institutional settings and often rely on informal networks for information and care [4].

The significant growth in FP counseling and method uptake suggests that the SKILLZ curriculum, when delivered within safe, youth-owned spaces, was successful in fostering both knowledge and motivation for behavior change. The social environment of the Youth Hubs appears to have served dual functions: as an access point for services and as a behavioral catalyst, normalizing SRH discussions and reducing stigma. The high conversion rate from non-clinical visits to FP adoption reinforces the importance of informal entry points, mentorship, games, peer-led sessions, as legitimate vehicles for health engagement.

Youth-friendly services, especially those embedded within peer-led, non-institutional settings, were central to behavior change observed in the program. Counseling, mentorship, and progressive exposure to FP information occurred in safe, familiar environments where adolescents did not feel judged or rushed. Many female participants, for instance, returned after one or two sessions before deciding to adopt a method, citing the need for time to “think through” their decision. Male adolescents more frequently requested condoms during initial visits, suggesting gendered preferences and norms in contraceptive behavior.

This echoes the findings of Sokol and Fisher [11], who stressed that autonomy, confidentiality, and relational safety are prerequisites for effective adolescent health programs. Similarly, Hailemariam et al. [10] found that bundling FP services within broader life development programming reduces stigma and improves outcomes, a design choice that YPE4AH explicitly followed. In Kano, where contraceptive conversations are highly sensitive, adolescents were more willing to engage with FP information when it was introduced through SKILLZ sessions focused on life goals, financial planning, and leadership. Future use was also established with a high intention to use established, a platform for service uptake when sexual debut happens for those not currently sexually active [28].

The strength of the hub model was further amplified by the program’s capacity to retain adolescents over time. Jakobsson et al. [1] noted that continuity of contact and community trust are critical enablers of SRH engagement. In YPE4AH, adolescent-friendly counseling and services were offered not as one-off interventions but as part of an ongoing ecosystem of support. This allowed adolescents to revisit their decisions, build confidence in service providers, and feel in control of their SRH journey.

Moreover, the impact estimates, over 7,000 unintended pregnancies averted and 39 maternal deaths prevented, highlight the value of investing in prevention and early engagement. These outcomes validate the design logic that embedded FP within a broader adolescent development framework. The multi-pronged approach was not merely additive; it was synergistic, positioning FP as one of several levers for adolescent agency, not a standalone intervention. Coupled with FP counseling and method uptake increasing significantly over time, the results suggest that the model was not only reaching more adolescents but doing so in a way that translated into real health outcomes.

By connecting safe spaces to a broader health ecosystem, YPE4AH created measurable health impact. Internally, the hubs contributed thousands of new FP acceptors, often starting with counseling before method initiation. Beyond the hubs, a total of 75,030 adolescents accessed FP through both hub-based and spoke referrals, illustrating how informal access points can anchor formal service uptake [4,5,10]. The diversity of methods (condoms, pills, injectables, implants, and even IUCDs) challenges the notion that adolescents will only accept short-term options, this aligns with Gottschalk et al. [19], who identified limited method mix and access options as critical gaps in adolescent SRH programming. Regional variations observed between northern and southern states (e.g., higher uptake of implants in the north, more condom and pill use in the south) highlight the importance of context-sensitive delivery strategies. By leveraging a multi-cadre referral system (including PHCs, private clinics, and community pharmacies) the program enabled adolescents to access a range of methods, including invasive ones like intrauterine contraceptive devices (IUCDs), without cost or stigma.

The sustained provision of free FP commodities through government collaboration enabled continuity of care, eliminating financial and psychological barriers for young people. This responds directly to Gottschalk’s call for long-term, systems-based programming, as well as to the need for equity in access [19]. It also validates the argument made by Kananura et al. that when adolescent-focused models are aligned with government systems and long-term resourcing (e.g., provision of free commodities through government channels), they become not just effective but sustainable [2]. The YPE4AH model provides an avenue to have both safe spaces as separate entities but still heavily linked with existing structure; a model that is worth replicating across the country and in other sub-Saharan African countries.

The impact modelling estimates (including unintended pregnancies and maternal deaths averted) demonstrate that even non-clinical entry points can be potent levers for public health outcomes when strategically embedded in community systems.

The broader literature often notes the challenge of reaching OSY, particularly in settings marked by poverty, stigma, or conflict [10, 29]. Yet, the YPE4AH project provides an evidence-based response to this challenge, combining service integration, youth ownership, and local partnerships to drive impact. This positions safe spaces not as fringe or pilot interventions, but as viable delivery platforms deserving of further scale.

Nonetheless, the paper acknowledges limitations. Exposure to all program components was not uniform, and the study design does not permit causal attribution of the combined approach to FP outcomes. Despite, the effect of the SKILLZ curriculum, the safe space environment and the provider network on health outcomes of participants, other factors may have contributed, which have not been explained through the longitudinal nature of the assessment that was used as part of the intervention. Taken as a whole, the ecosystem approach reflects the reality of how adolescents experience change, through layered reinforcing touchpoints.

The YPE4AH experience adds to the growing body of evidence that adolescent-responsive health systems must go beyond service availability to service approachability. For OSY, who often exist on the margins of formal systems, access must be relational, proximate, and affirming. Future efforts should explore how this model can be scaled or adapted to different urban and rural contexts, with stronger data systems to disaggregate impact across program strands.

## Conclusion

The YPE4AH project offers compelling evidence by demonstrating that for out-of-school adolescents (OSY) in urban slums, access to family planning can be meaningfully expanded through community-based safe spaces, youth-centered peer education within a multi-thronged approach, and a network of trusted providers. The results affirm that when adolescents are engaged through spaces they help shape, and supported by services that reflect their realities, health behaviors shift, stigma decreases, and uptake of critical services like FP increases; reaching marginalized populations and sustained over time.

While not all adolescents experienced the full range of program components, the foundational SKILLZ curriculum and the relational design of the Youth Hubs played a pivotal role in driving service engagement. Positioning FP within a broader frame of life skills and livelihood development helped anchor short-term health decisions in long-term personal aspirations.

Critically, what set this model apart was not simply its reach, but the way it redefined “integration.” Rather than relying on the conventional, and often ineffective strategy of embedding adolescent SRH into primary healthcare centers (PHCs), where concerns around privacy, stigma, and provider attitudes continue to deter young people, YPE4AH focused on relational integration. Safe spaces were designed to feel informal, judgment-free, and adolescent-owned. Yet, through close collaboration with government partners, including the coordinated provision of free FP commodities and the training of a multi-cadre referral network, these hubs remained connected to the broader health ecosystem without forcing adolescents into it.

This approach recognized the lived realities of OSY (many of whom actively avoid traditional institutions) and instead created parallel pathways that were trusted, accessible, and affirming. By making public sector resources visible and available within adolescent-friendly spaces, YPE4AH showed that effective integration is not about co-location of services, but about shared ownership, mutual reinforcement, and policy-backed collaboration.

While the study cannot disentangle the exact contribution of each program strand (life skills, livelihood and leadership), the cumulative outcomes, ranging from expanded method choice to thousands of new FP users and significant impact on adolescent health trajectories, affirm the promise of this ecosystem model. For countries seeking scalable, context-responsive solutions, YPE4AH presents a powerful blueprint; one that centers adolescent agency while strengthening public-private alignment in new, pragmatic ways.

## Data Availability

All data relating to the findings reported within the study are publicly available having been reported as part of an health intervention programming. Where required, the authors are willing to provide raw data to back the claims made in the manuscript.

## Acknowledgments

We would like to acknowledge and thank the Ministries of Health and Health Care Development Boards of Lagos and Kano states, including the Ministry of Youths of both states, Healthcare providers across both states who have shown commitment to providing quality services to adolescents, and have fully supported the YPE4AH implementation. We also like to acknowledge Hub coordinators, Youth Coaches and other staff that were involved in delivering the intervention that produced this case study. We also appreciate other members of the YPE4AH team in Lagos and Kano for their excellent work during the fieldwork and providing feedback on manuscript preparation.

The project from which this paper was culled was funded by the United States Agency for International Development (USAID), Agreement Number: 72062020CA00011. The opinions and inferences made in this work are those of the authors and not the funding agency.

## Supporting Information

## References

1. Jakobsson C, Sanghavi R, Nyamiobo J, Maloy C, Mwanzu A, Venturo-Conerly K, et al. Adolescent and youth-friendly health interventions in low-income and middle-income countries: a scoping review. BMJ Glob Health. 2024 Sep 5;9(9):e013393. doi:10.1136/bmjgh-2023-013393.

2. Kananura RM, Waiswa P, Melesse DY, Faye C, Boerma T. Examining the recent trends in adolescent sexual and reproductive health in five countries of sub-Saharan Africa based on PMA and DHS household surveys. Reprod Health. 2021 Jun 17;18(Suppl 1):121. doi:10.1186/s12978-021-01111-0.

3. Federal Ministry of Youth and Sports Development. National Youth Policy: enhancing youth development and participation in the context of sustainable development. 2019 ed. Abuja: Federal Ministry of Youth and Sports Development; 2019.

4. Adesola RO, Opuni E, Idris I, Okesanya OJ, Igwe O, Abdulazeez MD, et al. Navigating Nigeria’s health landscape: population growth and its health implications. Environ Health Insights. 2024 May 1;18:11786302241250211. doi:10.1177/11786302241250211.

5. Chandra-Mouli V, Greifinger R, Nwosu A, Hainsworth G, Sundaram L, Hadi S, et al. Invest in adolescents and young people: it pays. Reprod Health. 2013 Sep 16;10:51. doi:10.1186/1742-4755-10-51.

6. Loto OM, Ezechi OC, Kalu BK, Loto A, Ezechi L, Ogunniyi SO. Poor obstetric performance of teenagers: is it age- or quality of care-related? J Obstet Gynaecol. 2004 Jun;24(4):395–8. doi:10.1080/01443610410001685529.

7. Ebeigbe PN, Gharoro EP. Obstetric complications, intervention rates and maternofetal outcome in teenage nullipara in Benin City, Nigeria. Trop Doct. 2007 Apr;37(2):79–83. doi:10.1177/004947550703700206.

8. UNICEF. Evaluation report: the Out-of-School Children Initiative (OOSCI). New York: UNICEF; 2018.

9. YouthPower 2: Learning and Evaluation. Out-of-school youth [Internet]. Washington (DC): USAID; [cited 2024 Apr 20]. Available from: https://www.youthpower.org/youthpower-issues/topics/out-school-youth-0

10. Hailemariam S, Gutema L, Agegnehu W, Derese M. Challenges faced by female out-of-school adolescents in accessing and utilizing sexual and reproductive health service: a qualitative exploratory study in southwest Ethiopia. J Prim Care Community Health. 2021 Jan-Dec;12:21501327211018936. doi:10.1177/21501327211018936.

11. Sokol R, Fisher E. Peer support for the hardly reached: a systematic review. Am J Public Health. 2016;106(12):e1–8. doi:10.2105/AJPH.2016.303180.

12. Federal Ministry of Health (Nigeria). National policy on the health and development of adolescents and young people in Nigeria. Abuja: Federal Ministry of Health; 2007.

13. Federal Ministry of Health (Nigeria), Federal Ministry of Youth Development (Nigeria). Action plan for advancing young people’s health and development in Nigeria: 2010–2012. Abuja: Federal Ministry of Health and Federal Ministry of Youth Development; 2010.

14. Federal Ministry of Health (Nigeria). National guidelines for the integration of adolescent and youth friendly health services into primary healthcare facilities in Nigeria. Abuja: Federal Ministry of Health; 2013.

15. Federal Ministry of Health (Nigeria). Nigeria national standards and minimum service package for adolescent and youth-friendly health services. Abuja: Federal Ministry of Health; 2018.

16. Nigerian Educational Research and Development Council (NERDC). National family life and HIV education curriculum for junior secondary schools in Nigeria. Lagos: Federal Ministry of Education; 2003.

17. Grant MJ, Hallman KK. Pregnancy-related school dropout and prior school performance in KwaZulu-Natal, South Africa. Stud Fam Plann. 2008 Dec;39(4):369–82. doi:10.1111/j.1728-4465.2008.00181.x.

18. Brieger WR, Delano GE, Lane KG, Oladepo O, Oyediran KA. West African Youth Initiative: outcome of a reproductive health education program. J Adolesc Health. 2001 Nov;29(6):436–46. doi:10.1016/S1054-139X(01)00265-5.

19. Gottschalk LB, Ortayli N. Interventions to improve adolescents’ contraceptive behaviors in low- and middle-income countries: a review of the evidence base. Contraception. 2014 Sep;90(3):211–25. doi:10.1016/j.contraception.2014.04.017.

20. YouthPower. What young changemakers think of safe public spaces [Internet]. Washington (DC): USAID; 2018 [cited 2024 Apr 20]. Available from: https://www.youthpower.org/resources/what-young-changemakers-think-safe-public-spaces

21. Hershow R, Gannett K, Merrill J, Kaufman BE, Barkley C, DeCelles J, et al. Using soccer to build confidence and increase HCT uptake among adolescent girls: a mixed-methods study of an HIV prevention programme in South Africa. Sport Soc. 2015;18(8):1009–22. doi:10.1080/17430437.2014.997586.

22. Merrill KG, Merrill JC, Hershow RB, Barkley C, Rakosa B, DeCelles J, et al. Linking at-risk South African girls to sexual violence and reproductive health services: a mixed-methods assessment of a soccer-based HIV prevention program and pilot SMS campaign. Eval Program Plann. 2018;70:12–24. doi:10.1016/j.evalprogplan.2018.04.010.

23. Alvarado G, Skinner M, Plaut D, Moss C, Kapungu C, Reavley N. A systematic review of positive youth development programs in low- and middle-income countries. Washington (DC): YouthPower Learning, Making Cents International; 2017.

24. Ross DA, Hinton R, Melles-Brewer M, Engel D, Zeck W, Fagan L, et al. Adolescent well-being: a definition and conceptual framework. J Adolesc Health. 2020 Oct;67(4):472–6. doi:10.1016/j.jadohealth.2020.06.042.

25. Marie Stopes International (MSI). Impact 2 model [Internet]. London: MSI; 2023 Aug [cited 2024 Apr 20]. Available from: https://www.msichoices.org/what-we-do/technical-expertise/impact-2/

26. Villarruel AM, Zhou Y, Gallegos EC, Ronis DL. Examining long-term effects of Cuídate—a sexual risk reduction program in Mexican youth. Rev Panam Salud Publica. 2010 May;27(5):345–51. doi:10.1590/S1020-49892010000500004.

27. Erulkar AS, Muthengi E. Evaluation of Berhane Hewan: a program to delay child marriage in rural Ethiopia. Int Perspect Sex Reprod Health. 2009 Mar;35(1):6–14. doi:10.1363/ifpp.35.006.09.

28. Envuladu E, Agbo H, Ohize V, Zoakah A. Determinants and outcome of teenage pregnancy in a rural community in Jos, Plateau State, Nigeria. Sub-Saharan Afr J Med. 2014;1(1):48–54. doi:10.4103/2384-5147.129287.

29. Cartwright AF, Otai J, Maytan-Joneydi A, et al. Access to family planning for youth: perspectives of young family planning leaders from 40 countries. Gates Open Res. 2019;3:1513. doi:10.12688/gatesopenres.13045.2.

